# Defining the role of host biomarkers in the diagnosis and prognosis of childhood pneumonia – a prospective cohort study

**DOI:** 10.1101/2022.12.07.22283191

**Authors:** Arjun Chandna, Yoel Lubell, Lazaro Mwandigha, Phattaranit Tanunchai, Asama Vinitsorn, Melissa Richard-Greenblatt, Constantinos Koshiaris, Direk Limmathurotsakul, Francois Nosten, Mohammad Yazid Abdad, Rafael Perera-Salazar, Claudia Turner, Paul Turner

## Abstract

**Background:** Reliable tools to inform outpatient management of childhood pneumonia in resource-limited settings are needed. We investigated the value added by biomarkers of host infection response to the performance of the Liverpool quick Sequential Organ Failure Assessment score (LqSOFA), for triage of children presenting with pneumonia to a primary care clinic in a refugee camp on the Thailand-Myanmar border.

**Methods:** 900 presentations of children aged ≤ 24 months meeting WHO pneumonia criteria were included. The primary outcome was receipt of supplemental oxygen. We compared discrimination of a clinical risk score (LqSOFA) to markers of endothelial injury (Ang-1, Ang-2, sFlt-1), immune activation (CHI3L1, IP-10, IL-1ra, IL-6, IL-8, IL-10, sTNFR-1, sTREM-1), and inflammation (CRP, PCT), and quantified the net-benefit of including biomarkers alongside LqSOFA. We evaluated the differential contribution of LqSOFA and host biomarkers to the diagnosis and prognosis of severe pneumonia.

**Results:** 49/900 (5.4%) presentations met the primary outcome. Discrimination of LqSOFA and Ang-2, the best performing biomarker, were comparable (AUC 0.82 [95% CI 0.76-0.88] and 0.81 [95% CI 0.74-0.87] respectively). Combining Ang-2 with LqSOFA improved discrimination (AUC 0.91; 95% CI 0.87-0.94; p < 0.001), and resulted in greater net-benefit, with 10-30% fewer children requiring oxygen supplementation incorrectly identified as safe for community-based management. Ang-2 had greater prognostic utility than LqSOFA to identify children requiring supplemental oxygen later in their illness course.

**Conclusions:** Combining Ang-2 and LqSOFA could guide referrals of childhood pneumonia from resource-limited community settings. Further work on integration into patient triage is required.

## INTRODUCTION

Most cases of childhood pneumonia can be successfully managed at home.^1,2^ In remote locations of many low- and middle-income countries (LMICs), where accessing hospital-level care may incur substantial cost, community-based care is often preferred by families.^3^ However, pneumonia remains the leading cause of death and disability for young children living in LMICs and clear criteria for the safe outpatient management of pneumonia are required.^2,4,5^

The World Health Organization (WHO) recommends that primary care providers use the presence of ‘Danger Signs’ to determine whether a child with pneumonia requires referral to hospital.^6,7^ However, validity of these signs is unclear and they are affected by substantial interobserver reliability.^8-10^ In 2016, the quick Sequential Organ Failure Assessment (qSOFA) score was endorsed as a risk stratification tool for adults with suspected infection.^11^ Recently, an age-adapted version (the Liverpool-qSOFA [LqSOFA] score) was developed specifically for febrile children presenting from the community.^12^ In a recent analysis we demonstrated that the LqSOFA score outperformed two other paediatric severity scores in Southeast Asian children with acute respiratory infections (ARIs), suggesting that the score has excellent generalisability and may be practical for use in resource-limited primary care settings.^13^

A growing body of evidence indicates that final common pathophysiological pathways reflecting endothelial injury and immune activation are shared across a range of infectious diseases,^14,15^ including in young children with pneumonia.^16-18^ Markers of these pathways improve performance of clinical severity scores,^19^ including qSOFA,^15,20^ and consequently they have been proposed as adjuncts to paediatric triage tools.^21^ However, it is unknown whether such markers are elevated sufficiently early in the natural history of childhood pneumonia for them to be useful for risk stratification in primary care.

In this study we quantified the value that measurements of biomarkers of the host response to infection might add to clinical assessment to identify young children with pneumonia who are suitable for community-based management. We hypothesised that biomarker measurements would be most useful for prognostication in children not readily identified by clinical severity scores as requiring referral at the point of presentation.

## METHODS

### Study population

This was a secondary analysis of data collected during a prospective birth cohort study conducted between September 2007 and September 2010 on the Thailand-Myanmar border.^22^ Children of consenting pregnant women attending a medical clinic for refugees and internally-displaced people were reviewed at birth and followed-up each month (routine visit) and during any intercurrent illness (illness visit) until 24 months of age. The few other medical facilities and restricted movement out of the camp contributed to low attrition rates and enabled capture of the majority of acute illnesses for which care was sought. All illness visits meeting WHO pneumonia criteria (cough or difficulty breathing associated with age-adjusted tachypnoea) were included in this analysis.^23^

### Data collection

Clinical data were measured at presentation by the study team and entered on to structured case report forms. Heart rate and respiratory rate were measured over one minute. Mental status was assessed using the Alert Voice Pain Unresponsive (AVPU) scale. Capillary refill time was measured following the release of gentle pressure on the child’s sternum. Serum samples were collected in plain tubes at presentation. Participants were followed-up each day during admission to the clinic and at monthly routine visits conducted as part of the longitudinal birth cohort study.

### Primary and secondary outcomes

The primary outcome was receipt of supplemental oxygen during the illness visit, which was only indicated if peripheral oxygen saturation (SpO_2_) was < 90%. All staff were trained on the clinic treatment protocols prior to study commencement. To explore the diagnostic vs. prognostic value of the biomarkers, the secondary outcomes were SpO_2_ < 90% at enrolment (diagnostic outcome) and receipt of supplemental oxygen within 28 days of the illness visit (prognostic outcomes).

### Identification and selection of biomarkers

Host biomarkers were selected for analysis following review of the literature and expert consultation. A range of viral and bacterial pathogens commonly cause pneumonia in children and it is not possible to obtain a microbiological diagnosis in the vast majority of cases presenting to primary care. Acknowledging this and recognising therefore that clinically-useful biomarkers would need to be predictive across a spectrum of infecting organisms, we prioritised biomarkers implicated in ‘pathogen agnostic’ final common pathways to severe febrile illness and sepsis, including those reflecting endothelial injury (angiopoietin-1 [Ang-1], angiopoietin-2 [Ang-2], soluble fms-like tyrosine kinase-1 [sFlt-1; sVEGFR-1]) and immune activation (chitinase-3-like protein-1 [CHI3L1], interferon-gamma-inducible protein-10 [IP-10; CXCL-10], interleukin-1 receptor antagonist [IL-1ra], interleukin-6 [IL-6], interleukin-8 [IL-8], interleukin-10 [IL-10], soluble tumour necrosis factor receptor-1 [sTNFR-1], soluble triggering receptor expressed on myeloid cells-1 [sTREM-1]).^14-19,24,25^ We also included two acute phase proteins (C-reactive protein [CRP] and procalcitonin [PCT]); although previous studies have found them unhelpful for predicting the severity of childhood pneumonia,^26^ they are measurable using inexpensive commercially-available rapid tests and familiar to many clinicians.

### Laboratory procedures

Serum samples were centrifuged within two hours of collection (ambient temperature, at 3,000 rpm, for 10 minutes) and stored at 2-8°C. Each day, samples were transported using a cold-chain to the off-site laboratory, aliquoted, and stored at -80°C within 12 hours of collection. Samples collected on Saturday evening or Sunday were transported at the end of the working day on Monday (≤ 48 hours after collection). Frozen serum aliquots were thawed overnight and concentrations of host biomarkers were quantified using the Simple Plex Ella microfluidic platform (ProteinSimple, San Jose, California, USA).^27^ Analytes below the limit of quantification (LOQ) were assigned a value one-third of the lower limit of the standard curve (Supplementary Table 1).

### Missing data

Of the 900 included pneumonia presentations, 827 (91.9%; 827/900) had complete data for all baseline clinical predictor variables. Capillary refill time had the highest proportion of missingness (7.0%; 63/900; Supplementary Table 2). Median imputation grouped by outcome status was used to address missing observations.

### Statistical methods

A Lowess smoothing approach was used to explore the relationship between each biomarker and the primary outcome. We used univariable logistic regression to quantify the ability of the LqSOFA score and each individual biomarker to discriminate children presenting with pneumonia who required supplemental oxygen (area under the receiver operating characteristic curve [AUC]).

We compared the discrimination of the LqSOFA score to that of the LqSOFA score plus one biomarker (R package: *pROC*).^28,29^ To reduce the risk of multiple testing we limited comparisons to the five top-performing biomarkers, selected on the basis of their univariate discrimination, after confirming that none of the biomarker concentrations were strongly correlated with baseline LqSOFA scores (R package: *polycor*).^30^ We used decision curve analyses (R package: *dcurves*)^31^ to determine the net-benefit of including the biomarkers alongside the LqSOFA score and compared predicted classifications across a range of clinically-relevant referral thresholds.

To explore the differential contribution of biomarkers to the diagnosis and prognosis of severe pneumonia, we used univariable logistic regression to assess the ability of the LqSOFA score and the five top-performing biomarkers to discriminate children who were hypoxic at presentation (diagnostic outcome), and to discriminate children who were not initially hypoxic but whose disease progressed to require supplemental oxygen in the 28 days following presentation (prognostic outcomes).

Finally, recognising that a management strategy requiring measurement of a biomarker in every child presenting with pneumonia would not be practical, we used recursive partitioning to construct a proof-of-concept management algorithm combining the LqSOFA score and the top-performing biomarker to identify children who might be safe for community-based management. We acknowledged that safety and simplicity were paramount for community triage and specified a loss-matrix of 10:1 and maximum level of tree depth of two (R package: *rpart*).^32^

All analyses were conducted in R, version 4.0.2.^33^

### Sample size

No formal sample size calculation was performed for this secondary analysis. All available data were used to maximise power and generalisability of the results.

### Ethics and reporting

Ethical approvals were provided by the Mahidol University Ethics Committee (TMEC 21-023) and Oxford Tropical Research Ethics Committee (OxTREC 511-21). The study is reported in accordance with the Strengthening the Reporting of Observational Studies in Epidemiology (STROBE) guidelines (Supplementary Table 3).^34^

## RESULTS

Between September 2007 and September 2008, 999 pregnant women were enrolled and 965 children were born into the birth cohort. From September 2007 to September 2010 there were 900 presentations from 444 individual children which met the WHO criteria for pneumonia, had complete information about supplemental oxygen therapy, and had a serum sample available for analysis (Figure 1).

**FIGURE 1.**
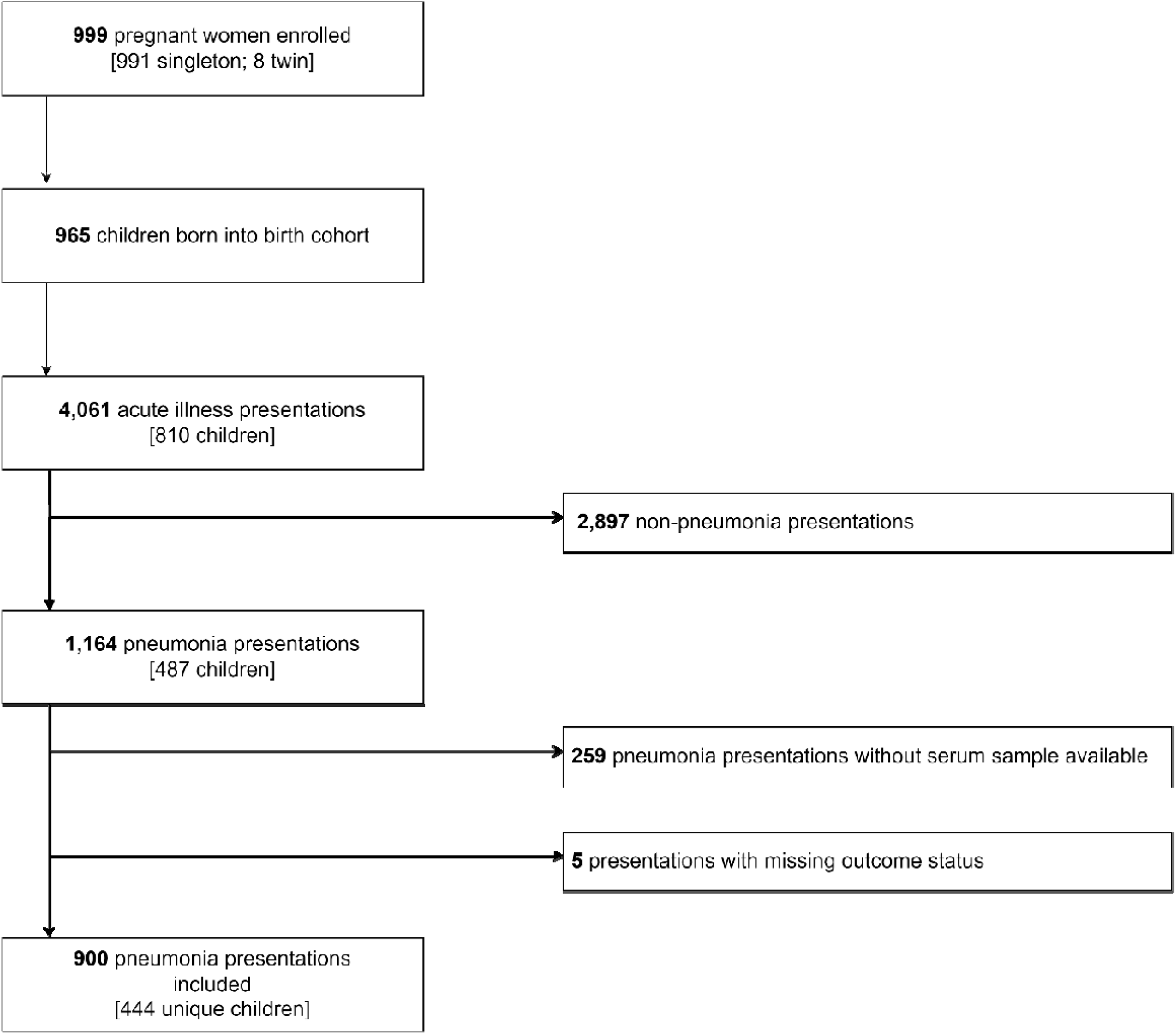
Eligibility of acute illness visits for inclusion in primary analysis.

Children had been symptomatic for a median of 3 days (interquartile range [IQR] 2 to 5 days) and fewer than 3% (2.8%; 25/900) had received antibiotics prior to enrolment (Table 1; Supplementary Table 4). Admission rate to the clinic was 28.4% (256/900). Forty-nine (5.4%; 49/900) presentations met the primary outcome. The LqSOFA score and biomarkers reflecting endothelial injury (Ang-2, sFlt-1), immune activation (IL-8, IL-6, IL-1ra), and inflammation (PCT) were associated with the primary outcome (Table 2; Supplementary Figure 1; p < 0.001).

**TABLE 1.** Baseline clinical characteristics of the cohort stratified by primary outcome status. ^a^Respiratory distress defined as head bobbing, tracheal tug, grunting and/or chest indrawing; ^b^abnormal chest auscultation defined as crepitations and/or wheeze; ^c^rectal temperature converted to axillary temperature for neonates and infants. ^†^Age-adjusted z-scores were calculated using the R package *z scorer*. For children admitted to the clinic, weight was measured at the time of presentation (seca scale; precision ± 5g for neonates or ± 50g after birth). All other anthropometric data were captured during routine visits and were eligible for inclusion in these analyses according to the following window periods: height ≤ 28 days; MUAC ≤ 28 days without intercurrent admission; weight ≤ 14 days without intercurrent admission. Median interval between anthropometric measurement and illness presentation: length = 8 days (IQR 4-12 days); MUAC = 9 days (IQR 4-13 days); weight = 4 days (IQR 0-9 days). ^*^Missing data: gestation = 3; birthweight = 7; comorbidity = 4; symptom duration = 3; abnormal lung auscultation = 13; lung crepitations = 15; wheeze = 22; heart rate = 2; respiratory rate = 1; temperature = 1; oxygen saturation = 138; capillary refill time = 63; mental status = 12; WLZ = 47; WAZ = 46; MAZ = 98; LAZ = 3.

| Characteristic | Overall<br>N = 900<sup>1</sup> | Supplemental oxygen |  | p-value<sup>2</sup> |
| --- | --- | --- | --- | --- |
|  |  | No<br>N = 851<sup>1</sup> | Yes<br>N = 49<sup>1</sup> |  |
| Demographics |  |  |  |  |
| Age (months) | 10.7 (6.0, 16.3) | 10.8 (6.0, 16.4) | 7.0 (3.4, 14.9) | 0.010 |
| Male sex | 456 / 900 (51%) | 432 / 851 (51%) | 24 / 49 (49%) | 0.80 |
| Birth history |  |  |  |  |
| Gestation (weeks)* | 39.1 (38.1, 40.0) | 39.1 (38.1, 40.0) | 38.5 (37.3, 39.5) | 0.011 |
| Birthweight (kg)* | 2.9 (2.6, 3.2) | 2.9 (2.6, 3.2) | 2.6 (2.0, 3.1) | 0.006 |
|  |  | No<br>N = 851<sup>1</sup> | Yes<br>N = 49<sup>1</sup> |  |
| Previous medical history |  |  |  |  |
| Number of previous illness visits | 4.0 (2.0, 7.0) | 4.0 (2.0, 7.0) | 3.0 (2.0, 8.0) | 0.40 |
| Time since last illness visit (days) | 45.0 (15.0, 106.2) | 47.0 (17.0, 109.0) | 18.0 (1.0, 69.0) | 0.002 |
| Known comorbidity<sup>*</sup> | 11 / 896 (1.2%) | 7 / 849 (0.8%) | 4 / 47 (8.5%) | 0.002 |
| History of current illness |  |  |  |  |
| Duration of symptoms (days)<sup>*</sup> | 3.0 (2.0, 5.0) | 3.0 (2.0, 5.0) | 3.0 (2.0, 5.0) | 0.060 |
| Antibiotics prior to presentation | 25 / 900 (2.8%) | 24 / 851 (2.8%) | 1 / 49 (2.0%) | >0.90 |
| Presenting symptoms and signs |  |  |  |  |
| Fever | 780 / 900 (87%) | 743 / 851 (87%) | 37 / 49 (76%) | 0.018 |
| Cough | 895 / 900 (99%) | 846 / 851 (99%) | 49 / 49 (100%) | >0.90 |
| Respiratory distress<sup>a</sup> | 278 / 900 (31%) | 233 / 851 (27%) | 45 / 49 (92%) | <0.001 |
| Head bobbing | 33 / 900 (3.7%) | 15 / 851 (1.8%) | 18 / 49 (37%) | <0.001 |
| Tracheal tug | 76 / 900 (8.4%) | 53 / 851 (6.2%) | 23 / 49 (47%) | <0.001 |
| Grunting | 14 / 900 (1.6%) | 5 / 851 (0.6%) | 9 / 49 (18%) | <0.001 |
| Chest indrawing | 273 / 900 (30%) | 228 / 851 (27%) | 45 / 49 (92%) | <0.001 |
|  |  | No<br>N = 851<sup>1</sup> | Yes<br>N = 49<sup>1</sup> |  |
| Abnormal lung auscultation<sup>b*</sup> | 719 / 887 (81%) | 676 / 839 (81%) | 43 / 48 (90%) | 0.12 |
| Crepitations<sup>*</sup> | 659 / 885 (74%) | 621 / 838 (74%) | 38 / 47 (81%) | 0.30 |
| Wheeze<sup>*</sup> | 338 / 878 (38%) | 311 / 830 (37%) | 27 / 48 (56%) | 0.009 |
| **Vital signs** |  |  |  |  |
| Heart rate (bpm)<sup>*</sup> |  |  |  |  |
| Neonate | 149.0 (143.8, 160.0) | 154.0 (147.2, 160.0) | 145.0 (140.0, 153.5) | 0.70 |
| Infant | 140.0 (130.0, 148.0) | 140.0 (130.0, 148.0) | 144.0 (134.0, 158.0) | 0.019 |
| Child | 132.0 (124.0, 140.0) | 132.0 (124.0, 140.0) | 136.0 (128.0, 141.5) | 0.20 |
| Respiratory rate (bpm)<sup>*</sup> |  |  |  |  |
| Neonate | 67.0 (63.5, 77.0) | 63.0 (61.0, 65.5) | 78.0 (73.0, 80.5) | 0.081 |
| Infant | 58.0 (54.0, 60.8) | 58.0 (54.0, 60.0) | 60.0 (56.0, 64.0) | 0.036 |
| Child | 50.0 (46.0, 56.0) | 50.0 (46.0, 56.0) | 58.0 (46.5, 61.5) | 0.046 |
| Axillary temperature (°C)<sup>c*</sup> | 37.1 (36.4, 37.7) | 37.1 (36.4, 37.7) | 37.3 (36.5, 38.1) | 0.13 |
| Oxygen saturation (%)<sup>*</sup> | 95.0 (93.0, 96.0) | 95.0 (93.0, 96.0) | 88.0 (86.0, 92.0) | <0.001 |
| Capillary refill time > 2 secs<sup>*</sup> | 12 / 837 (1.4%) | 8 / 792 (1.0%) | 4 / 45 (8.9%) | 0.003 |
|  |  | No<br>N = 851<sup>1</sup> | Yes<br>N = 49<sup>1</sup> |  |
| Not alert<sup>*</sup> | 116 / 888 (13%) | 87 / 842 (10%) | 29 / 46 (63%) | <0.001 |
| **Anthropometrics** |  |  |  |  |
| Weight-for-length z-score (WLZ)<sup>*†</sup> | -0.2 (-0.9, 0.6) | -0.2 (-0.9, 0.6) | -0.4 (-1.7, 0.7) | 0.067 |
| Weight-for-age z-score (WAZ)<sup>*†</sup> | -1.0 (-1.9, -0.4) | -1.0 (-1.8, -0.4) | -1.9 (-3.4, -0.4) | <0.001 |
| MUAC-for-age z-score (MAZ)<sup>*†</sup> | 0.1 (-0.6, 0.7) | 0.1 (-0.5, 0.7) | -1.2 (-2.0, -0.2) | <0.001 |
| Length-for-age z-score (LAZ)<sup>*†</sup> | -1.6 (-2.5, -0.9) | -1.6 (-2.4, -0.8) | -2.5 (-3.0, -1.3) | 0.003 |

<sup>1</sup>Median (IQR); n / N (%)
<sup>2</sup>Wilcoxon rank sum test; Pearson's Chi-squared test; Fisher's exact test

**TABLE 2.** Baseline LqSOFA scores and host biomarker concentrations of the cohort, stratified by primary outcome status.

|  | Overall<br>N = 900<sup>1</sup> | Supplemental oxygen |  | p-value<sup>2</sup> |
| --- | --- | --- | --- | --- |
|  |  | No<br>N = 851<sup>1</sup> | Yes<br>N = 49<sup>1</sup> |  |
| LqSOFA score |  |  |  |  |
| 0 | 713 / 900 (79%) | 703 / 851 (83%) | 10 / 49 (20%) | <0.001 |
| 1 | 156 / 900 (17%) | 128 / 851 (15%) | 28 / 49 (57%) |  |
| 2 | 30 / 900 (3.3%) | 20 / 851 (2.4%) | 10 / 49 (20%) |  |
| 3 | 1 / 900 (0.1%) | 0 / 851 (0%) | 1 / 49 (2.0%) |  |
| Host biomarkers |  |  |  |  |
| Endothelial injury |  |  |  |  |
| Ang-1 (pg/ml) | 33,913.5 (21,938.0, 46,836.5) | 33,897.0 (21,849.0, 46,572.0) | 35,880.0 (23,177.0, 50,341.0) | 0.50 |
| Ang-2 (pg/ml) | 1,538.5 (1,187.0, 1,984.8) | 1,485.0 (1,164.5, 1,915.5) | 2,261.0 (1,792.0, 3,412.0) | <0.001 |
| sFlt-1 (pg/ml) | 170.0 (145.0, 196.0) | 168.0 (144.0, 193.0) | 197.0 (171.0, 231.0) | <0.001 |
| Immune activation |  |  |  |  |
| CHI3L1 (ng/ml) | 43.2 (31.0, 61.4) | 43.2 (31.1, 60.8) | 44.0 (30.0, 67.8) | 0.70 |
| IL-1ra (pg/ml) | 1,929.5 (1,217.0, 2,949.8) | 1,890.0 (1,210.5, 2,794.0) | 3,519.0 (1,858.0, 5,136.0) | <0.001 |
| IL-6 (pg/ml) | 17.4 (8.9, 36.4) | 16.9 (8.7, 34.6) | 38.9 (13.8, 70.7) | <0.001 |
|  |  | No<br>N = 851<sup>1</sup> | Yes<br>N = 49<sup>1</sup> |  |
| IL-8 (pg/ml) | 35.8 (23.8, 52.0) | 34.9 (23.1, 51.0) | 53.7 (37.0, 82.1) | <0.001 |
| IL-10 (pg/ml) | 17.0 (11.2, 28.0) | 16.9 (11.0, 27.2) | 22.5 (13.5, 38.7) | 0.015 |
| IP-10 (pg/ml) | 745.0 (441.0, 1,371.0) | 742.0 (438.5, 1,350.5) | 886.0 (551.0, 1,834.0) | 0.064 |
| sTNFR-1 (pg/ml) | 1,583.5 (1,345.8, 1,906.0) | 1,575.0 (1,341.0, 1,894.0) | 1,821.0 (1,545.0, 2,196.0) | 0.001 |
| sTREM-1 (pg/ml) | 399.5 (315.0, 522.0) | 398.0 (313.5, 520.5) | 423.0 (345.0, 533.0) | 0.20 |
| *Acute phase proteins* |  |  |  |  |
| CRP (mg/L) | 20.4 (7.4, 44.9) | 20.1 (7.2, 43.8) | 31.0 (7.5, 67.9) | 0.20 |
| PCT (pg/ml) | 240.0 (176.0, 391.2) | 236.0 (173.0, 372.5) | 417.0 (248.0, 722.0) | <0.001 |

<sup>1</sup>Median (IQR); n / N (%)
<sup>2</sup>Wilcoxon rank sum test; Pearson's Chi-squared test; Fisher's exact test

### Ang-2, sFlt-1, and IL-8 improve discrimination of the LqSOFA score

Ang-2 demonstrated substantially better discrimination than any other biomarker and comparable discrimination to the LqSOFA score (Table 3). No biomarker outperformed the clinical LqSOFA score (Supplementary Table 5). The relationships between baseline biomarker concentrations and the probability of supplemental oxygen requirement are shown (Supplementary Figure 2).

**TABLE 3.** Ability of the LqSOFA score and host biomarkers to discriminate children who required supplemental oxygen, and the value added to the clinical LqSOFA score by the five top-performing biomarkers.

| Predictor | AUC (95% CI) |  | p-value<sup>1</sup> |
| --- | --- | --- | --- |
|  | Univariate | + LqSOFA |  |
| **LqSOFA** | – | 0.82 (0.76-0.88) | – |
| **Ang-2** | 0.81 (0.74-0.87) | 0.91 (0.87-0.94) | < 0.001 |
| **IL-8** | 0.72 (0.65-0.79) | 0.88 (0.85-0.92) | 0.001 |
| **sFlt-1** | 0.69 (0.61-0.77) | 0.89 (0.86-0.92) | < 0.001 |
| **PCT** | 0.69 (0.62-0.77) | 0.78 (0.69-0.86) | 0.015 |
| **IL-1ra** | 0.68 (0.59-0.77) | 0.80 (0.72-0.88) | 0.242 |
| **IL-6** | 0.65 (0.56-0.74) |  |  |
| **sTNFR-1** | 0.64 (0.55-0.72) |  |  |
| **IL-10** | 0.60 (0.52-0.69) |  |  |
| **IP-10** | 0.58 (0.49-0.66) |  |  |
| **sTREM-1** | 0.56 (0.49-0.63) |  |  |
| **CRP** | 0.55 (0.46-0.64) |  |  |
| **Ang-1** | 0.53 (0.44-0.62) |  |  |
| **CHI3L1** | 0.52 (0.43-0.61) |  |  |

<sup>1</sup>DeLong method to compare AUC of LqSOFA vs. AUC of LqSOFA + biomarker.

As baseline LqSOFA scores and biomarker concentrations were not strongly correlated (Supplementary Tables 6-7; Supplementary Figure 3) we selected the five most discriminatory biomarkers and quantified the improvement in discrimination of the LqSOFA score when combined with each biomarker in turn. Discrimination improved when either Ang-2, sFlt-1 or IL-8 were combined with LqSOFA (Table 3; p < 0.001 to 0.001).

### Combining Ang-2 and LqSOFA improves identification of children suitable for community-based management of pneumonia

We recognised that better discrimination does not necessarily translate into greater utility and acknowledged that the relative value of a true negative (correctly identifying a child who could be safely managed in the community) and a false negative (misclassifying a child who would require supplemental oxygen) is context-dependent.^35,36^ We used decision curve analyses to account for this and compared the net-benefit of the LqSOFA score alone to that of the LqSOFA score combined with either Ang-2, sFlt-1 or IL-8, over a range of clinically-plausible referral thresholds.^37^ At referral thresholds beyond ∼8% (a management strategy equivalent to referring any child in whom the predicted risk of requiring supplemental oxygen is ≥ 8%) addition of Ang-2 to the LqSOFA score provided greater utility than the LqSOFA score alone (Figure 2). Examining predicted classifications across these referral thresholds suggested that an algorithm combining Ang-2 and LqSOFA could reduce the number of children incorrectly identified as safe for community-based management by ∼10-30% compared to the LqSOFA score alone, without substantially increasing the proportion of unnecessary referrals (Table 4). Addition of neither sFlt-1 nor IL-8 provided greater net-benefit than the LqSOFA score alone at any referral threshold (Figure 2).

**TABLE 4.** Predicted classifications at different referral thresholds using the LqSOFA score and the LqSOFA score combined with Ang-2. A referral threshold of 5% reflects a management strategy whereby any child with a predicted probability of requiring oxygen ≥ 5% is referred. ^*^LqSOFA scores converted to predicted probabilities to facilitate comparison with the LqSOFA score + Ang-2; referral thresholds (predicted probabilities) approximate to the following LqSOFA scores: 1% ≈ ≥ 0; 5% ≈ ≥ 1; 20% ≈ ≥ 2; 40% ≈ ≥ 3.

| Referral threshold | Sensitivity (95% CI) | Specificity (95% CI) | Negative Predictive Value (95% CI) | Positive Predictive Value (95% CI) | Negative Likelihood Ratio (95% CI) | Positive Likelihood Ratio (95% CI) | Cases referred (%) | Cases managed in community (%) | Ratio of Incorrect to Correct referrals | Ratio of Correct to Incorrect cases managed in community |
| --- | --- | --- | --- | --- | --- | --- | --- | --- | --- | --- |
| **LqSOFA score\*** |  |  |  |  |  |  |  |  |  |  |
| **1%** | 1.00 (NA) | 0.00 (NA) | 1.00 (NA) | 0.05 (0.04-0.07) | 0.00 (NA) | 1.00 (NA) | 900 (100%) | 0 (0%) | 17 to 1 | NA |
| **5%** | 0.79 (0.67-0.89) | 0.83 (0.80-0.85) | 0.99 (0.98-0.99) | 0.21 (0.15-0.26) | 0.25 (0.13-0.40) | 4.56 (3.63-5.54) | 184 (20.4%) | 716 (79.6%) | 4 to 1 | 71 to 1 |
| **20%** | 0.23 (0.13-0.35) | 0.98 (0.97-0.99) | 0.96 (0.94-0.97) | 0.35 (0.18-0.52) | 0.79 (0.67-0.90) | 9.95 (4.62-16.91) | 28 (3.1%) | 872 (96.9%) | 2 to 1 | 22 to 1 |
| **40%** | 0.20 (0.00-0.33) | 0.98 (0.97-1.00) | 0.96 (0.94-0.97) | 0.43 (0.31-1.00) | 0.82 (0.68-1.00) | Inf (NA) | 1 (0.1%) | 899 (99.9%) | 0 to 1 | 18 to 1 |
| **LqSOFA score + Ang-2** |  |  |  |  |  |  |  |  |  |  |
| **1%** | 1.00 (0.91-1.00) | 0.20 (0.00-0.59) | 1.00 (1.00-1.00) | 0.07 (0.05-0.11) | 0.00 (0.00-0.13) | 1.32 (1.00-2.66) | 559 (62.1%) | 341 (37.9%) | 10 to 1 | Inf to 1 |
| **5%** | 0.85 (0.69-0.93) | 0.82 (0.76-0.87) | 0.99 (0.98-0.99) | 0.21 (0.17-0.26) | 0.19 (0.09-0.37) | 4.73 (3.74-7.50) | 202 (22.4%) | 698 (77.6%) | 4 to 1 | 77 to 1 |
| **20%** | 0.43 (0.27-0.56) | 0.96 (0.95-0.97) | 0.97 (0.95-0.98) | 0.40 (0.28-0.50) | 0.59 (0.45-0.76) | 11.73 (7.26-19.28) | 58 (6.4%) | 842 (93.6%) | 2 to 1 | 23 to 1 |
| **40%** | 0.29 (0.08-0.46) | 0.99 (0.97-0.99) | 0.96 (0.95-0.97) | 0.52 (0.28-0.67) | 0.72 (0.55-0.93) | 21.28 (7.56-37.98) | 22 (2.4%) | 878 (97.6%) | 1 to 1 | 23 to 1 |

**FIGURE 2.**
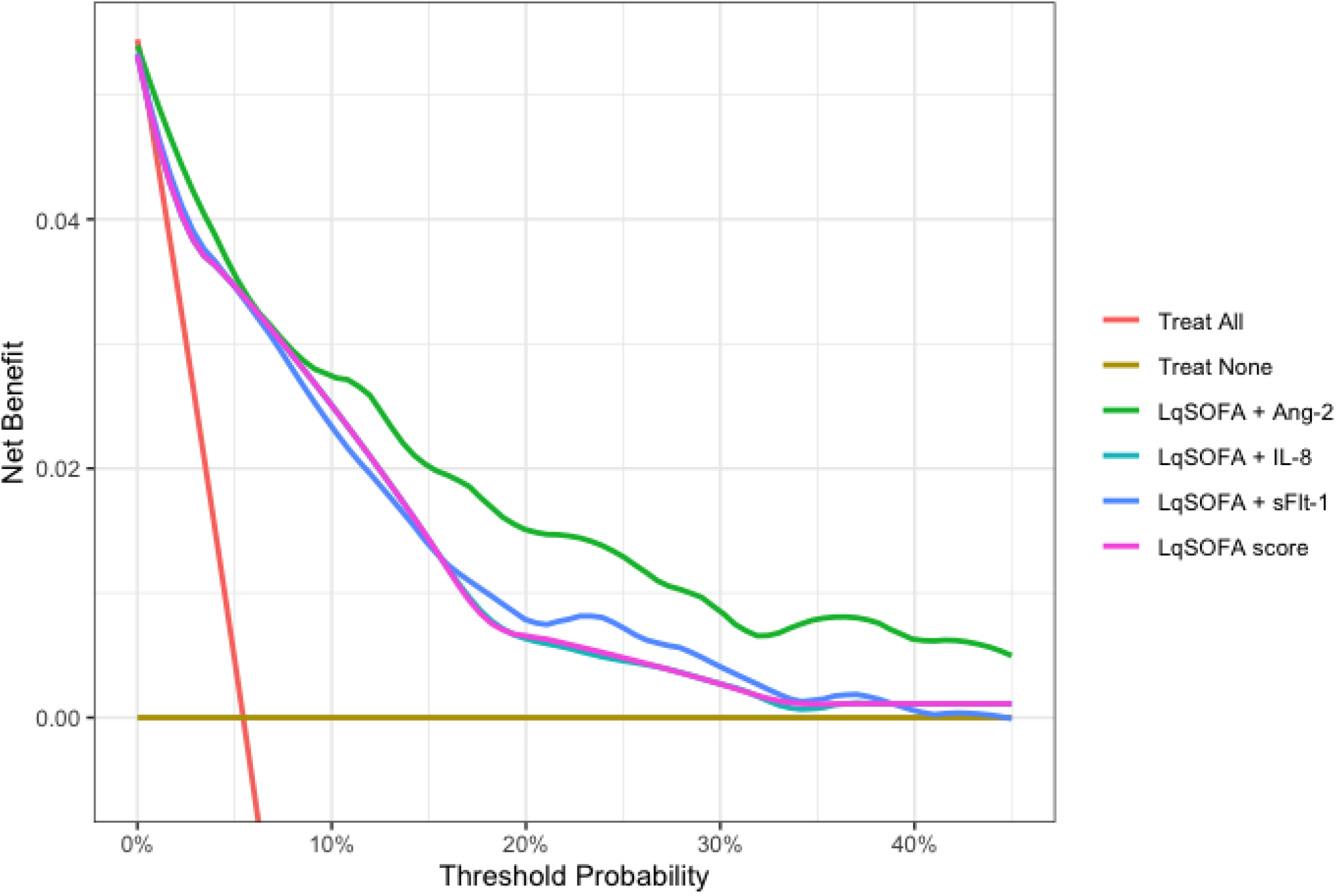
Utility of the LqSOFA score alone and combined with Ang-2, sFlt-1 or IL-8 for the identification of children suitable for community-based management of pneumonia. The net-benefit of the LqSOFA score (pink line) is compared to the LqSOFA score combined with either Ang-2 (green line), IL-8 (turquoise line), or sFlt-1 (blue line), and a “refer-all” (red line) and “refer-none” (brown line) approach. A threshold probability of 5% is equivalent to a management strategy in which any child with a predicted risk of supplemental oxygen requirement ≥ 5% is referred (i.e., a scenario where the value of one correct referral is equivalent to 19 incorrect referrals or a number-needed-to-refer of 20).

### Differing roles for host biomarkers in the diagnosis and prognosis of severe pneumonia

To explore the differing contributions of host biomarkers to the diagnosis and prognosis of severe pneumonia we examined the ability of the five most discriminatory biomarkers and LqSOFA score to distinguish children who were: (a) hypoxic at enrolment (SpO_2_ < 90%); (b) not hypoxic at enrolment but required supplemental oxygen during their illness visit; and (c) not hypoxic at enrolment but required supplemental oxygen at any time in the 28 days following enrolment. Discrimination of the LqSOFA score deteriorated for the more prognostic outcomes, whereas discrimination of the host biomarkers appeared stable (Table 5). Decision curve analyses confirmed Ang-2 to have greater prognostic utility (net-benefit) than either IL-8 or the LqSOFA score (Supplementary Figure 4).

**TABLE 5.** Discrimination of the LqSOFA score and host biomarkers for the diagnosis and prognosis of severe pneumonia. Children with hypoxia (SpO_2_ < 90%) at presentation excluded for assessment of prognostic outcomes. Presentations with missing outcome status excluded: Outcome 1 assessed in 766 presentations (734 controls and 32 cases); Outcome 2 assessed in 869 presentations (846 controls and 23 cases); Outcome 3 assessed in 827 presentations (789 cases and 38 cases).

| OUTCOME 1 (diagnosis)<br>SpO<sub>2</sub> < 90% at enrolment |  | OUTCOME 2 (prognosis)<br>Supplemental O<sub>2</sub> during illness visit |  | OUTCOME 3 (prognosis)<br>Supplemental O<sub>2</sub> ≤ 28 days after enrolment |  |
| --- | --- | --- | --- | --- | --- |
| Predictor | AUC (95% CI) | Predictor | AUC (95% CI) | Predictor | AUC (95% CI) |
| LqSOFA | 0.85 (0.78-0.91) | Ang-2 | 0.80 (0.70-0.90) | IL-8 | 0.75 (0.67-0.82) |
| Ang-2 | 0.74 (0.65-0.84) | IL-8 | 0.75 (0.64-0.85) | Ang-2 | 0.73 (0.64-0.82) |
| sFlt-1 | 0.71 (0.62-0.81) | LqSOFA | 0.75 (0.64-0.85) | IL-1ra | 0.70 (0.61-0.80) |
| IL-1ra | 0.71 (0.61-0.81) | PCT | 0.68 (0.56-0.80) | PCT | 0.67 (0.59-0.76) |
| IL-8 | 0.70 (0.62-0.77) | IL-1ra | 0.66 (0.53-0.79) | LqSOFA | 0.66 (0.57-0.74) |
| PCT | 0.69 (0.59-0.78) | sFlt-1 | 0.60 (0.47-0.73) | sFlt-1 | 0.65 (0.56-0.75) |

### An algorithm for the safe outpatient management of childhood pneumonia

We combined the LqSOFA score and Ang-2 to generate a simple proof-of-concept algorithm for triage of all children presenting with pneumonia (Figure 3). Since sensitivity would usually be prioritised for community-based triage, we specified the cost of misclassifying a child who would require supplemental oxygen as 10 times that of the cost of misclassifying a child who would not, reflecting a pragmatic approximation for the upper limit of the number-needed-to-refer (NNR; number of children referred in order to identify one child who would progress to require supplemental oxygen) from a typical resource-limited primary care setting. The algorithm achieved a negative likelihood ratio (NLR) of 0.28 (sensitivity = 78.1%) and positive likelihood ratio (PLR) of 3.66 (specificity = 78.7%), for the identification of children suitable for home-based management of pneumonia.

**FIGURE 3.**
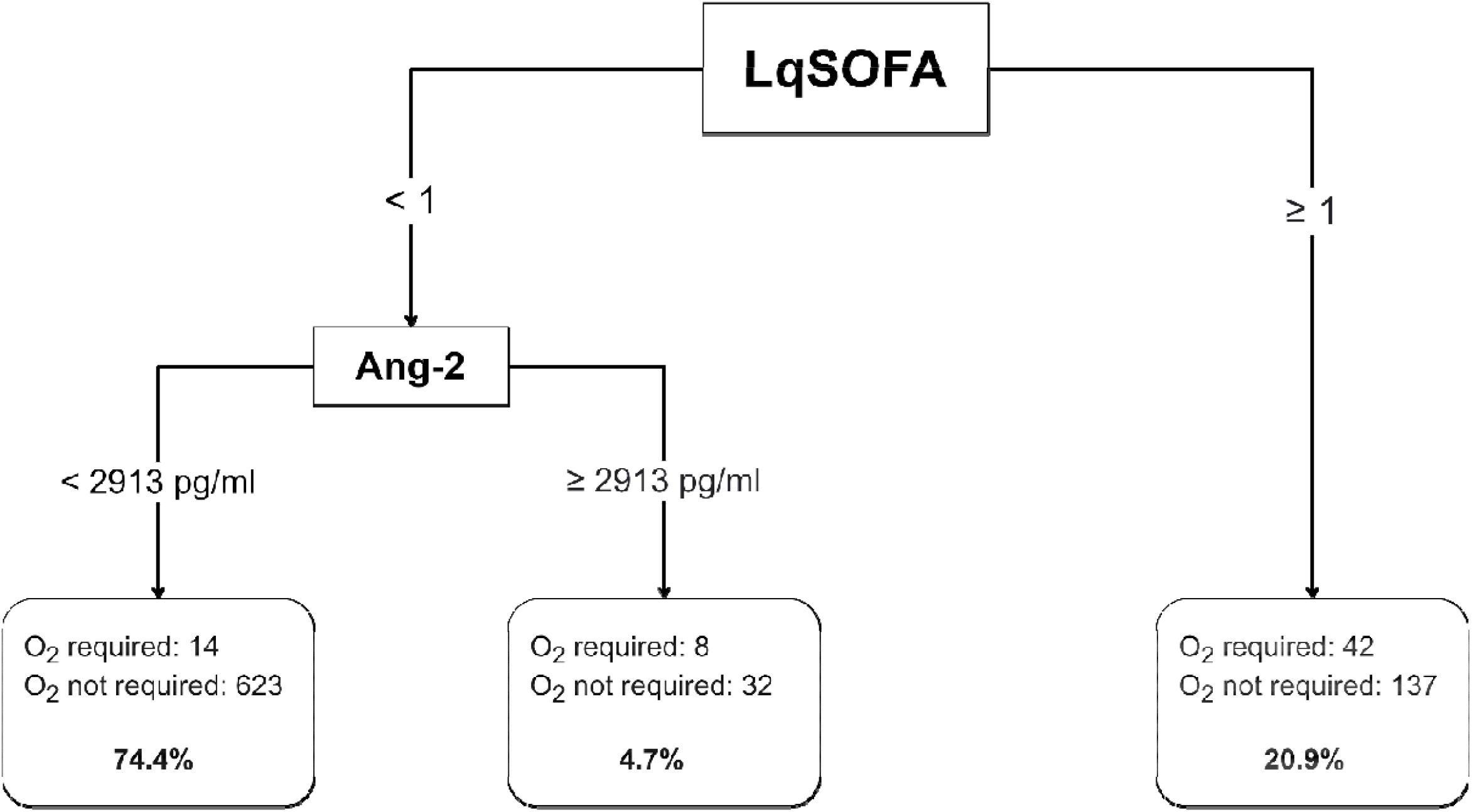
Triage of childhood pneumonia in resource-limited primary care contexts. The cost of misclassifying a child who would require supplemental oxygen was prespecified as 10 times that of misclassifying a child who would not require supplemental oxygen. Cut-points selected by recursive partitioning. Percentages indicate proportion of cohort in each terminal node. O_2_ = oxygen.

## DISCUSSION

We report the promising performance of Ang-2, a marker of endothelial injury, for risk stratification of young children presenting with pneumonia to a primary care clinic located within a refugee camp on the Thailand-Myanmar border. Combining the LqSOFA score with Ang-2 improved sensitivity of the LqSOFA score alone and resulted in safer identification of children suitable for community-based management across a range of clinically-plausible referral thresholds. Furthermore, amongst children not hypoxic at presentation, baseline Ang-2 concentrations were able to identify those whose illnesses subsequently progressed over the next 28 days, outperforming other biomarkers and the LqSOFA score.

The performance of the LqSOFA score is consistent with our broader analysis of the score in children with ARIs,^13^ and comparable to that reported in the original derivation and validation study.^12^ The LqSOFA score is the largest age-adapted version of the widely-endorsed qSOFA score for adults,^38^ and was specifically designed for triaging children presenting from the community. Unlike other paediatric pneumonia risk scores it uses routinely collected data, which facilitated external validation in our resource-limited primary care setting.^39^

Multiple univariate analyses can emphasise chance findings and hence we elected not to compare the performance of individual biomarkers to the LqSOFA score in the main analysis. Policy makers and healthcare workers are most likely to adopt biomarker tests as adjuncts to clinical risk stratification, although they have been proposed as standalone replacements in settings with limited capacity for collection of clinical data.^40^ In our cohort, whilst Ang-2 and LqSOFA had comparable discrimination, the net-benefit of the LqSOFA score appeared superior (Supplementary Table 8; Supplementary Figure 5).

Our results illustrate the critical importance of considering clinical context when evaluating potential incremental value of biomarkers, rather than relying on summary measures such as the AUC alone.^36^ Although some biomarkers of endothelial injury (Ang-2, sFlt-1) and immune activation (IL-8) improved the ability of the LqSOFA score to discriminate children who required supplemental oxygen, only for Ang-2 did this translate into superior clinical utility (net-benefit).

Including measurements of Ang-2 alongside the LqSOFA score could make triage of paediatric pneumonia safer. Sensitivity improved such that ∼10-30% fewer children would be incorrectly identified for community-based management, without increasing the proportion of inappropriate referrals. However, laboratory tests carry an opportunity cost, especially in settings with limited resources, and it should be noted that this strategy would require the measurement of Ang-2 in all children presenting with pneumonia. Whether this is feasible would depend on many factors, including the availability, durability, turnaround time, and cost of a point-of-care test for Ang-2. Should such a test become available cost-effectiveness analyses accounting for differing scenarios would be required before it could be recommended for use.

An alternative strategy, perhaps more compatible with the clinical workflow and resources available in busy LMIC primary care settings, could be to use the easily practicable LqSOFA score as a screening tool to identify high-risk children with pneumonia, and measure Ang-2 concentrations only in the remaining subset of children not readily identified as requiring referral to hospital by the LqSOFA score. Using this approach, we were able to achieve a sensitivity of 78.1% (50/64) for identifying children who would require supplemental oxygen over the 28 days following presentation, whilst maintaining an incorrect-to-correct referral ratio of 3:1 (i.e., an NNR of four; specificity of 78.7%), and reducing the number of Ang-2 tests performed by more than 20% (179/856). Further efficiencies could be achieved by converting the points-based LqSOFA score into a clinical prediction model, which would permit the identification of both low- and high-risk groups who could be adequately risk stratified without measurement of Ang-2.

The association between higher concentrations of Ang-2 and supplemental oxygen requirement has biological plausibility. Ang-2 destabilises the endothelium, increases microvascular permeability, and is implicated in the pathogenesis of acute lung injury and sepsis.^14,19,24,41^ Previous work has illustrated the prognostic role of Ang-2 in adults with pneumonia and in hospitalised children with hypoxaemic pneumonia.^16,19^ Although this is the first study to investigate the role of Ang-2 in paediatric pneumonia at the community-level, endothelial dysfunction has been documented in ambulatory children with mild ARIs.^42^

Recently, the immune activation marker sTREM-1 has been shown to be prognostic in hospitalised children with pneumonia and proposed as a possible risk stratification tool.^17,18^ In our cohort, baseline sTREM-1 concentrations were similar in children who did and did not progress to require supplemental oxygen. As McDonald et al. note, the results of hospital-based studies cannot be generalised to community settings; in our study most children (71.6%; 644/900) were managed in the community and only a quarter (26.2%; 236/900) had severe pneumonia at presentation, compared to over three-quarters of children who had severe pneumonia at the time sTREM-1 levels were measured in previous hospital-based studies.^17,18^ Furthermore, studies of adults with Covid-19 suggest that sTREM-1 may be useful for predicting mortality but less well-suited for predicting proximal outcomes such as supplemental oxygen requirement.^43,44^

This is the largest study investigating the role of markers of endothelial injury and immune activation in paediatric pneumonia, and the only study to our knowledge conducted at the community-level. The local circumstances of our cohort enabled us to recruit children at the first point of contact with the formal health system and follow-up those managed as outpatients, aspects that are critical for robust evaluation of clinical scores and biomarkers in primary care.^9^ We evaluated a pre-specified panel of biomarkers with mechanistic links to severe respiratory disease and quantified the value they might add to a validated clinical score. We adopted an analytical approach which acknowledged that the threshold for home-based management of pneumonia would vary in different healthcare settings and that there is an inherent difference between recognising a child who is acutely unwell at the time of presentation and identifying a child who appears clinically stable but is at risk of subsequent deterioration.^45^

Our secondary analysis was limited to the data that were available. Presentations without serum samples were excluded. These presentations were more likely to have respiratory distress, altered mental status, and receive supplemental oxygen (Supplementary Table 9), and thus future studies should assess whether our findings are generalisable to more severe pneumonia presentations in the community. For the secondary (diagnostic) outcome, 15.3% (139/905) of presentations were excluded as baseline SpO_2_ measurements were missing.

Missingness is unlikely to be at random as measurement of SpO_2_ was a prerequisite for children considered for supplemental oxygen therapy: 89.2% (124/139) of missing values were in outpatients and no presentations missing baseline SpO_2_ received supplemental oxygen. A sensitivity analysis assuming that all presentations missing baseline SpO_2_ measurements were not hypoxic (SpO_2_ ≥ 90%) produced almost identical results (Supplementary Table 10).

The results of the recursive partitioning analysis will inherently reflect the 10:1 trade-off between false negatives and false positives specified in the loss-matrix. Whilst this was informed by our clinical experience of working in resource-limited settings, and is comparable to approaches taken by other groups,^20,46^ it will not apply in all contexts. We were unable to cross-validate our decision trees and hence the results will be optimistic and should be viewed as indicative of a framework within which Ang-2 and LqSOFA might be jointly deployed for triage of childhood pneumonia at the community level.

Samples collected at weekends were stored at 2-8°C for > 12 hours prior to being transferred to -80°C. Although most biomarkers are stable at refrigeration temperatures for short periods following centrifugation,^47^ we performed a sensitivity analysis excluding weekend presentations, which produced similar results (Supplementary Table 11).

Simple pathogen agnostic algorithms could be particularly impactful in resource-limited primary care settings where patient management is often syndromic and the infecting pathogen is usually unknown at the time of initial assessment. We demonstrate that measurements of Ang-2, a biomarker of endothelial injury, could improve the sensitivity of a validated clinical score and may enable safer community-based triage of childhood pneumonia. Combinatorial approaches integrating clinical risk scores and host biomarker measurements could assist health workers identify children who are acutely unwell at presentation and those who will deteriorate later, enabling earlier and more appropriate referrals to higher-level care. Future prospective work should focus on validating our findings and investigate the clinical utility and cost-effectiveness of different strategies for integrating biomarker measurements into patient assessment and triage.

## Data Availability

De-identified, individual participant data from this study will be available to researchers whose proposed purpose of use is approved by the data access committees at the Mahidol-Oxford Tropical Medicine Research Unit. Inquiries or requests for the data may be sent to.

## ABBREVIATIONS

ANG-1: angiopoietin-1
ANG-2: angiopoietin-2
ARI: acute respiratory infection
AUC: area under the receiver operating characteristic curve
AVPU: Alert Voice Pain Unresponsive
CHI3L1: chitinase-3-like protein-1
CI: confidence interval
CRP: C-reactive protein
IL-1ra: interleukin-1 receptor antagonist
IL-6: interleukin-6
IL-8: interleukin-8
IL-10: interleukin-10
IP-10: inducible protein-10 [CXCL-10]
IQR: interquartile range
LAZ: length-for-age z-score
LMIC: low- and middle-income country
LOQ: limit of quantification
LqSOFA: Liverpool quick Sequential Organ Failure Assessment
MAZ: MUAC-for-age z-score
MUAC: mid-upper arm circumference
NLR: negative likelihood ratio
OxTREC: Oxford Tropical Research Ethics Committee
PCT: procalcitonin
PLR: positive likelihood ratio
qSOFA: quick Sequential Organ Failure Assessment
sFlt-1: soluble fms-like tyrosine kinase-1 [sVEGFR-1]
SpO_2_: 75 peripheral oxygen saturation
sTNFR-1: soluble tumour necrosis factor receptor-1
sTREM-1: soluble triggering receptor expressed on myeloid cells-1
STROBE: Strengthening the Reporting of Observational Studies in Epidemiology
TMEC: Tropical Medicine Ethics Committee
USA: United States of America
WAZ: weight-for-age z-score
WLZ: weight-for-length z-score
WHO: World Health Organization

