## Appendix for "Defining the role of host biomarkers in the diagnosis and prognosis of childhood pneumonia – a prospective cohort study"

### SUPPLEMENTARY APPENDIX

**CONTENTS** **PAGE**


| | | |
|---|---|---|
| 1. | **Analytes below the limit of quantification** | 2 |
| 2. | **Missing data for constituent variables in the LqSOFA score** | 3 |
| 3. | **STROBE checklist** | 4 |
| 4. | **Baseline characteristics for clinical patient-level variables** | 6 |
| 5. | **Relationship between baseline LqSOFA score and observed outcome proportions** | 7 |
| 6. | **Discrimination of the LqSOFA score and individual host biomarkers** | 8 |
| 7. | **Relationships between baseline biomarker concentrations and primary outcome status** | 9 |
| 8. | **Relationships between baseline biomarker concentrations and baseline LqSOFA score I** | 10 |
| 9. | **Relationships between baseline biomarker concentrations and baseline LqSOFA score II** | 12 |
| 10. | **Correlation between baseline biomarker concentrations and baseline LqSOFA score** | 13 |
| 11. | **Prognostic clinical utility of Ang-2, IL-8 and LqSOFA for children with pneumonia** | 14 |
| 12. | **Predicted classifications at different referral thresholds using Ang-2 or LqSOFA** | 15 |
| 13. | **Net-benefit of Ang-2 and LqSOFA, alone and in combination, for triage of childhood pneumonia** | 16 |
| 14. | **Comparison of presentations with and without a serum sample available for biomarker analysis** | 17 |
| 15. | **Sensitivity analysis assuming all presentations with missing baseline $SpO_2$ are not hypoxic** | 20 |
| 16. | **Exclusion of samples which may have been stored at 2-8°C for > 12 hours prior to freezing** | 21 |

**Supplementary Table 1. Analytes below the limit of quantification.** Analytes below the limit of quantification were assigned a value one-third of the lower limit of the standard curve. Ang-1 = angiopoietin-1; CHI3L1 = chitinase-3-like protein-1; CRP = C-reactive protein; IL-6 = interleukin-6; IL-8 = interleukin-8; IL-10 = interleukin-10; sFlt-1 = soluble fms-like tyrosine kinase-1; sTNFR-1 = soluble tumour necrosis factor receptor-1.

| Biomarker | Number below the limit of quantification |
|---|---|
| **Ang-1** | 2 (0·2%) |
| **CHI3L1** | 4 (0·4%) |
| **CRP** | 13 (1·4%) |
| **IL-10** | 11 (1·2%) |
| **IL-6** | 7 (0·8%) |
| **IL-8** | 1 (0·1%) |
| **sTNFR-1** | 2 (0·2%) |
| **sFlt-1** | 5 (0·6%) |

**Supplementary Table 2. Missing data for constituent variables in the LqSOFA score.** LqSOFA = Liverpool quick Sequential Organ Failure Assessment.

| Variables | Number of missing data |
|---|---|
| **Heart rate** | 2 (0·2%) |
| **Respiratory rate** | 1 (0·1%) |
| **Capillary refill time** | 63 (7·0%) |
| **Mental status** | 12 (1·3%) |

**Supplementary Table 3. STROBE checklist.**

| | Item No | Recommendation | Page No |
|---|---|---|---|
| **Title and abstract** | 1 | (*a*) Indicate the study's design with a commonly used term in the title or the abstract | 4 |
| | | (*b*) Provide in the abstract an informative and balanced summary of what was done and what was found | 4 |
| **Introduction** | | | |
| Background/rationale | 2 | Explain the scientific background and rationale for the investigation being reported | 5-6 |
| Objectives | 3 | State specific objectives, including any prespecified hypotheses | 6 |
| **Methods** | | | |
| Study design | 4 | Present key elements of study design early in the paper | 6-7 |
| Setting | 5 | Describe the setting, locations, and relevant dates, including periods of recruitment, exposure, follow-up, and data collection | 6-7 |
| Participants | 6 | (*a*) Give the eligibility criteria, and the sources and methods of selection of participants. Describe methods of follow-up<br>(*b*) For matched studies, give matching criteria and number of exposed and unexposed | 6-7 |
| Variables | 7 | Clearly define all outcomes, exposures, predictors, potential confounders, and effect modifiers. Give diagnostic criteria, if applicable | 7 |
| Data sources/ measurement | 8* | For each variable of interest, give sources of data and details of methods of assessment (measurement). Describe comparability of assessment methods if there is more than one group | 7-8 |
| Bias | 9 | Describe any efforts to address potential sources of bias | 10 |
| Study size | 10 | Explain how the study size was arrived at | 10 |
| Quantitative variables | 11 | Explain how quantitative variables were handled in the analyses. If applicable, describe which groupings were chosen and why | 9 |
| Statistical methods | 12 | (*a*) Describe all statistical methods, including those used to control for confounding<br>(*b*) Describe any methods used to examine subgroups and interactions<br>(*c*) Explain how missing data were addressed<br>(*d*) If applicable, explain how loss to follow-up was addressed<br>(*e*) Describe any sensitivity analyses | 9-10 |
| **Results** | | | |
| Participants | 13* | (a) Report numbers of individuals at each stage of study—eg numbers potentially eligible, examined for eligibility, confirmed eligible, included in the study, completing follow-up, and analysed<br>(b) Give reasons for non-participation at each stage<br>(c) Consider use of a flow diagram | 11<br>Fig 1 |
| Descriptive data | 14* | (a) Give characteristics of study participants (eg demographic, clinical, social) and information on exposures and potential confounders<br>(b) Indicate number of participants with missing data for each variable of interest<br>(c) Summarise follow-up time (eg, average and total amount) | 11<br>Table 1<br>Table 2<br>Table S1<br>Table S2 |
| Outcome data | 15* | Report numbers of outcome events or summary measures over time | 11 |

| | | | |
|---|---|---|---|
| Main results | 16 | (*a*) Give unadjusted estimates and, if applicable, confounder-adjusted estimates and their precision (eg, 95% confidence interval). Make clear which confounders were adjusted for and why they were included<br>(*b*) Report category boundaries when continuous variables were categorized<br>(*c*) If relevant, consider translating estimates of relative risk into absolute risk for a meaningful time period | 12-14 |
| Other analyses | 17 | Report other analyses done—eg analyses of subgroups and interactions, and sensitivity analyses | 17-18 |
| **Discussion** | | | |
| Key results | 18 | Summarise key results with reference to study objectives | 14 |
| Limitations | 19 | Discuss limitations of the study, taking into account sources of potential bias or imprecision. Discuss both direction and magnitude of any potential bias | 17-18 |
| Interpretation | 20 | Give a cautious overall interpretation of results considering objectives, limitations, multiplicity of analyses, results from similar studies, and other relevant evidence | 18 |
| Generalisability | 21 | Discuss the generalisability (external validity) of the study results | 14-16 |
| **Other information** | | | |
| Funding | 22 | Give the source of funding and the role of the funders for the present study and, if applicable, for the original study on which the present article is based | 1-2 |

*Give information separately for exposed and unexposed groups.

**Note:** An Explanation and Elaboration article discusses each checklist item and gives methodological background and published examples of transparent reporting. The STROBE checklist is best used in conjunction with this article (freely available on the Web sites of PLoS Medicine at http://www.plosmedicine.org/, Annals of Internal Medicine at http://www.annals.org/, and Epidemiology at http://www.epidem.com/). Information on the STROBE Initiative is available at http://www.strobe-statement.org.

**Supplementary Table 4. Baseline characteristics of the cohort for patient-level variables, stratified by outcome status.** *Missing data: gestation = 2; birthweight = 4.

| Characteristic | Overall N = 444[1] | Supplemental oxygen | | p-value[2] |
|---|---|---|---|---|
| | | No N = 418[1] | Yes N = 26[1] | |
| Male sex | 220 / 444 (50%) | 205 / 418 (50%) | 15 / 26 (58%) | 0·40 |
| Gestation (weeks)* | 39·1 (38·2, 40·0) | 39·1 (38·2, 39·6) | 39·2 (38·1, 40·2) | 0·80 |
| Birthweight (kg)* | 2·9 (2·6, 3·2) | 2·9 (2·6, 3·2) | 2·9 (2·6, 3·2) | 0·80 |

[1]Median (IQR); n / N (%)

[2]Wilcoxon rank sum test; Pearson's Chi-squared test; Fisher's exact test

**Supplementary Figure 1. Relationship between baseline LqSOFA score and observed outcome proportions.** LqSOFA = Liverpool quick Sequential Organ Failure Assessment.

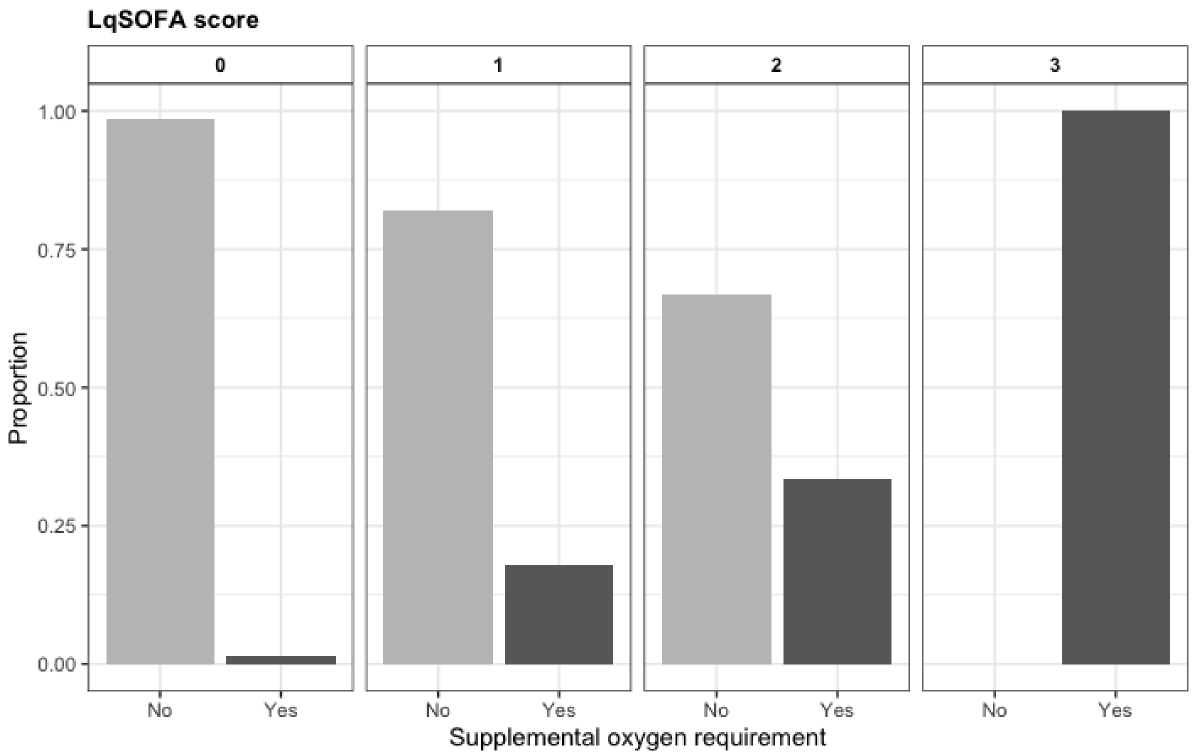

**Supplementary Table 5. Ability of the LqSOFA score and individual host biomarkers to discriminate children who required supplemental oxygen.** As a biomarker would most likely be used to supplement clinical assessment (rather than in isolation), and because univariate comparisons can emphasise chance associations, we chose not to perform statistical tests to compare the performance of individual biomarkers and LqSOFA score in the main analysis. We provide them here for interested readers.

| Predictor | **AUC** (95% CI) | p-value[1] |
|---|---|---|
| **LqSOFA** | 0·82 (0·76-0·88) | – |
| **Ang-2** | 0·81 (0·74-0·87) | 0·74 |
| **IL-8** | 0·72 (0·65-0·79) | 0·04 |
| **sFlt-1** | 0·69 (0·61-0·77) | 0·02 |
| **PCT** | 0·69 (0·62-0·77) | 0·01 |
| **IL-1ra** | 0·68 (0·59-0·77) | 0·004 |
| **IL-6** | 0·65 (0·56-0·74) | 0·001 |
| **sTNFR-1** | 0·64 (0·55-0·72) | 0·001 |
| **IL-10** | 0·60 (0·52-0·69) | < 0·001 |
| **IP-10** | 0·58 (0·49-0·66) | < 0·001 |
| **sTREM-1** | 0·56 (0·49-0·63) | < 0·001 |
| **CRP** | 0·55 (0·46-0·64) | < 0·001 |
| **Ang-1** | 0·53 (0·44-0·62) | < 0·001 |
| **CHI3L1** | 0·52 (0·43-0·61) | < 0·001 |

[1]DeLong method to compare AUC of LqSOFA vs. AUC of biomarker.

**Supplementary Figure 2. Relationship between baseline biomarker concentrations and probability of supplemental oxygen requirement.** Biomarker concentrations plotted on Log2 scale**.** Black line = probability of oxygen requirement; grey shaded area = 95% confidence interval. Ang-1 = angiopoietin-1; Ang-2 = angiopoietin-2; CHI3L1 = chitinase-3-like protein-1; CRP = C-reactive protein; IL-1ra = interleukin-1 receptor antagonist; IL-6 = interleukin-6; IL-8 = interleukin-8; IL-10 = interleukin-10; IP-10 = interferon-gamma-inducible protein-10; IQR = interquartile range; $O_2$ = oxygen; PCT = procalcitonin; sFlt-1 = soluble fms-like tyrosine kinase-1; sTNFR-1 = soluble tumour necrosis factor receptor-1; sTREM-1 = soluble triggering receptor expressed on myeloid cells-1.

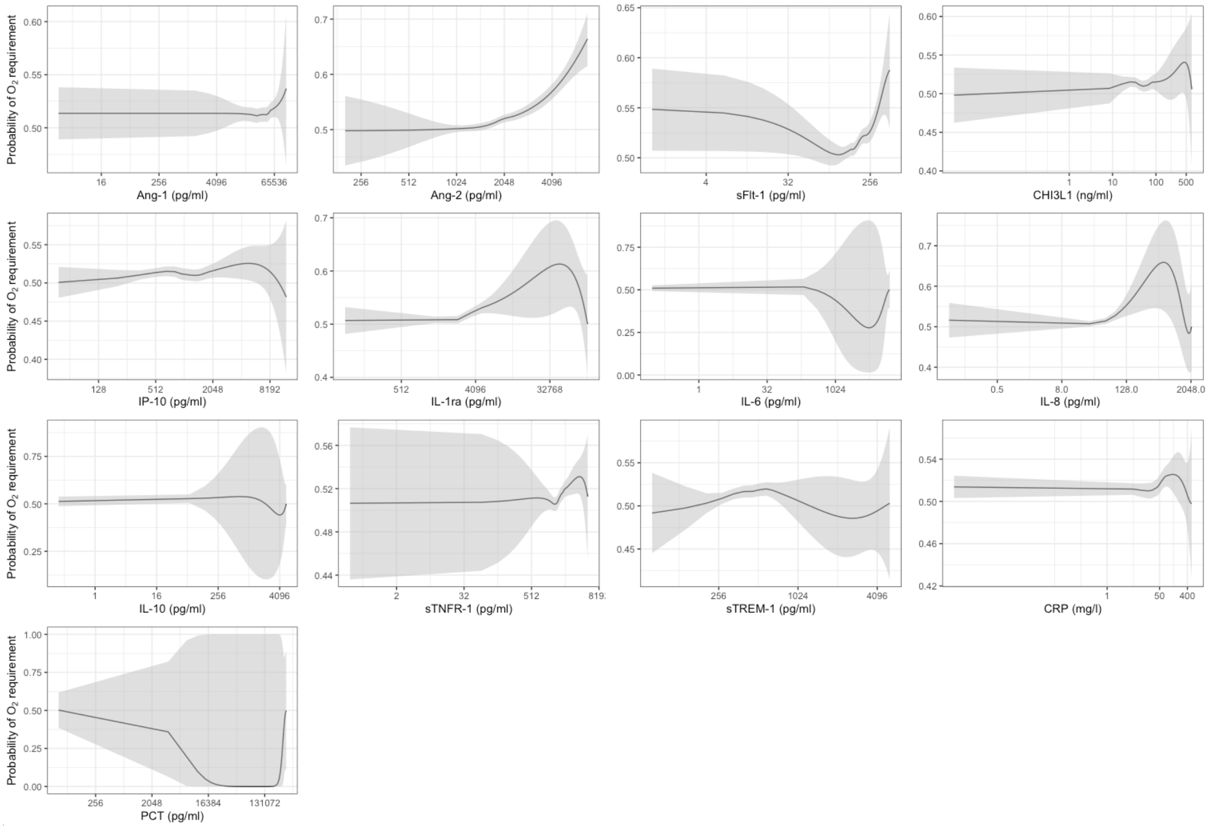

**Supplementary Table 6. Baseline biomarker concentrations stratified by level of baseline LqSOFA score.** Ang-1 = angiopoietin-1; Ang-2 = angiopoietin-2; CHI3L1 = chitinase-3-like protein-1; CRP = C-reactive protein; IL-1ra = interleukin-1 receptor antagonist; IL-6 = interleukin-6; IL-8 = interleukin-8; IL-10 = interleukin-10; IP-10 = interferon-gamma-inducible protein-10; IQR = interquartile range; LqSOFA = Liverpool quick Sequential Organ Failure Assessment; PCT = procalcitonin; sFlt-1 = soluble fms-like tyrosine kinase-1; sTNFR-1 = soluble tumour necrosis factor receptor-1; sTREM-1 = soluble triggering receptor expressed on myeloid cells-1.

| Biomarker | LqSOFA score | | | |
|---|---|---|---|---|
| | 0<br>N = 713[1] | 1<br>N = 156[1] | 2<br>N = 30[1] | 3<br>N = 1 |
| *Endothelial injury* | | | | |
| Ang1 (pg/ml) | 34,661·0 (22,376·0, 47,143·0) | 30,963·0 (21,157·5, 43,494·0) | 39,321·5 (29,893·8, 63,535·2) | 16,686·0 |
| Ang2 (pg/ml) | 1,480·0 (1,150·0, 1,889·0) | 1,703·5 (1,288·5, 2,241·2) | 2,031·0 (1,592·8, 2,782·0) | 3,412·0 |
| sFlt-1 (pg/ml) | 166·0 (143·0, 190·0) | 174·5 (150·0, 205·2) | 197·5 (177·2, 243·2) | 337·0 |
| *Immune activation* | | | | |
| CHI3L1 (ng/ml) | 43·0 (30·9, 59·8) | 42·6 (29·3, 60·8) | 61·5 (40·8, 99·5) | 208·3 |
| IL-1ra (pg/ml) | 1,807·0 (1,177·0, 2,673·0) | 2,601·0 (1,497·5, 4,113·5) | 4,109·0 (2,242·2, 6,739·0) | 59,522·0 |
| IL-6 (pg/ml) | 15·8 (8·3, 31·4) | 26·5 (11·2, 49·9) | 57·0 (13·9, 105·5) | 180·0 |
| IL-8 (pg/ml) | 33·5 (22·2, 49·0) | 43·5 (29·8, 58·8) | 53·4 (35·1, 97·4) | 72·2 |
| IL-10 (pg/ml) | 16·4 (10·6, 26·6) | 19·8 (13·5, 29·2) | 20·9 (13·9, 36·8) | 232·0 |
| IP-10 (pg/ml) | 706·0 (425·0, 1,289·0) | 937·0 (557·8, 1,492·0) | 1,398·0 (754·0, 1,882·0) | 4,081·0 |
| sTNFR-1 (pg/ml) | 1,557·0 (1,340·0, 1,865·0) | 1,682·5 (1,340·5, 2,048·2) | 1,868·0 (1,617·8, 2,233·8) | 4,555·0 |
| sTREM-1 (pg/ml) | 390·0 (310·0, 509·0) | 427·0 (326·5, 530·2) | 550·0 (426·5, 675·8) | 652·0 |
| *Acute phase proteins* | | | | |
| CRP (mg/l) | 18·4 (6·7, 40·0) | 27·9 (10·2, 56·5) | 40·5 (13·5, 85·1) | 186·0 |

| Biomarker | LqSOFA score | | | |
|---|---|---|---|---|
| | 0<br>N = 713[1] | 1<br>N = 156[1] | 2<br>N = 30[1] | 3<br>N = 1 |
| PCT (pg/ml) | 222·0 (168·0, 350·0) | 301·0 (208·2, 694·5) | 460·5 (258·2, 1,342·5) | 19,459·0 |

[1]Median (IQR)

**Supplementary Figure 3. Baseline biomarker concentrations stratified by level of baseline LqSOFA score.** Log2 scales used to plot concentrations in pg/ml (Ang-1, Ang-2, IL-1ra, IL-6, Il-8, IL-10, IP-10, PCT, sFlt-1, sTNFR-1, sTREM-1), ng/ml (CHI3L1), and mg/L (CRP). Ang-1 = angiopoietin-1; Ang-2 = angiopoietin-2; CHI3L1 = chitinase-3-like protein-1; CRP = C-reactive protein; IL-1ra = interleukin-1 receptor antagonist; IL-6 = interleukin-6; IL-8 = interleukin-8; IL-10 = interleukin-10; IP-10 = interferon-gamma-inducible protein-10; LqSOFA = Liverpool quick Sequential Organ Failure Assessment; PCT = procalcitonin; sFlt-1 = soluble fms-like tyrosine kinase-1; sTNFR-1 = soluble tumour necrosis factor receptor-1; sTREM-1 = soluble triggering receptor expressed on myeloid cells-1.

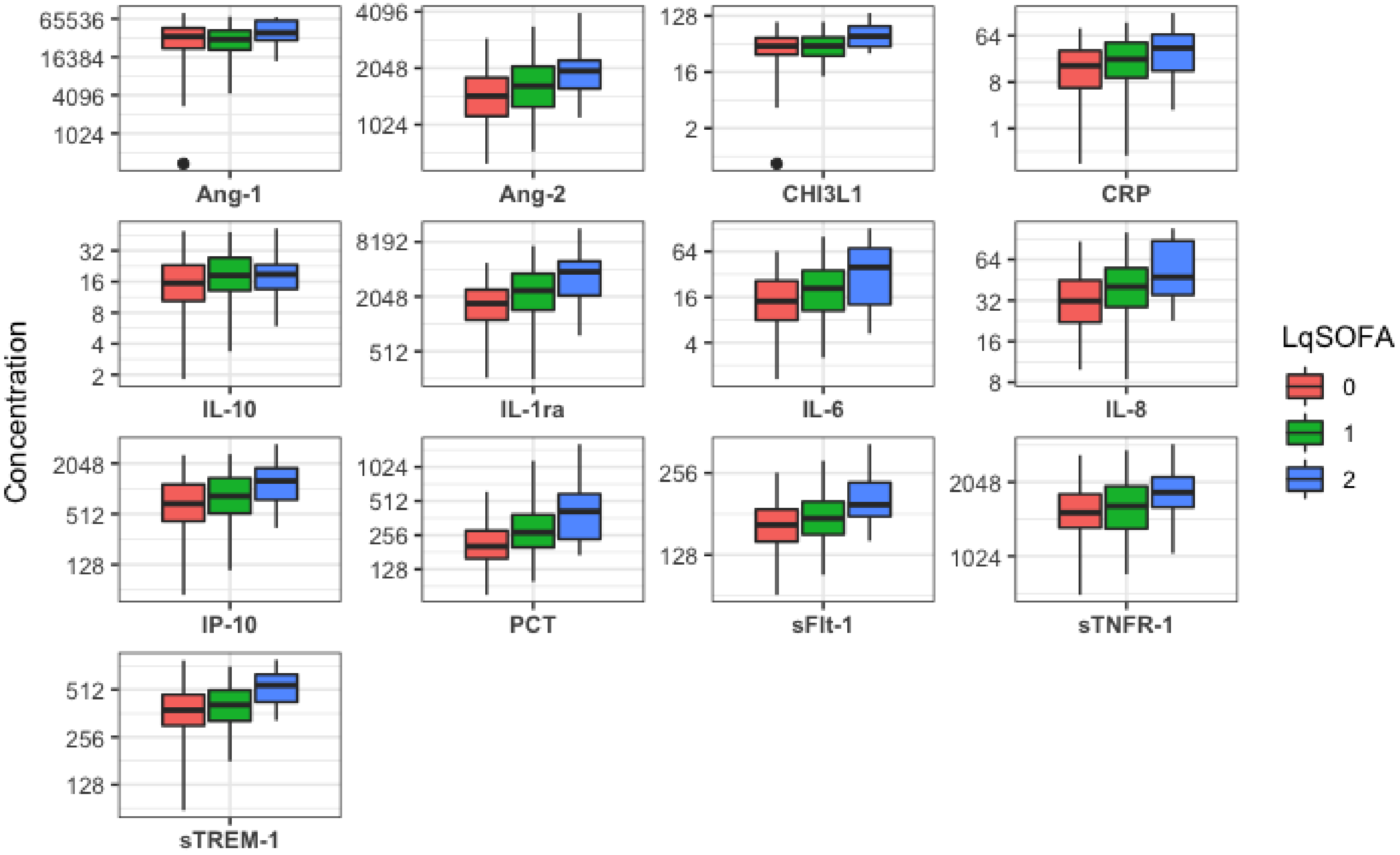

**Supplementary Table 7. Correlation between baseline biomarker concentrations and baseline LqSOFA scores.** Ang-1 = angiopoietin-1; Ang-2 = angiopoietin-2; CHI3L1 = chitinase-3-like protein-1; CRP = C-reactive protein; IL-1ra = interleukin-1 receptor antagonist; IL-6 = interleukin-6; IL-8 = interleukin-8; IL-10 = interleukin-10; IP-10 = interferon-gamma-inducible protein-10; LqSOFA = Liverpool quick Sequential Organ Failure Assessment; PCT = procalcitonin; sFlt-1 = soluble fms-like tyrosine kinase-1; sTNFR-1 = soluble tumour necrosis factor receptor-1; sTREM-1 = soluble triggering receptor expressed on myeloid cells-1.

| Biomarker | Polyserial correlation coefficient |
|---|---|
| *Endothelial injury* | |
| Ang-1 | 0·007 |
| Ang-2 | 0·322 |
| sFlt-1 | 0·280 |
| *Immune activation* | |
| CHI3L1 | 0·187 |
| IL-1ra | 0·409 |
| IL-6 | 0·145 |
| IL-8 | 0·177 |
| IL-10 | 0·093 |
| IP-10 | 0·210 |
| sTNFR-1 | 0·279 |
| sTREM-1 | 0·065 |
| *Acute phase proteins* | |
| PCT | 0·163 |
| CRP | 0·196 |

**Supplementary Figure 4. Prognostic utility of Ang-2, IL-8 and the LqSOFA score to predict supplemental oxygen requirement within the next 28 days for children with pneumonia.** The net-benefit of the LqSOFA score (pink line) is compared to Ang-2 (green line) or IL-8 (blue line), and a "refer-all" (red line) and "refer-none" (brown line) approach. A threshold probability of 5% is equivalent to a management strategy in which any child with a predicted risk of supplemental oxygen requirement ≥ 5% is referred (i.e., a scenario where the value of one correct referral is equivalent to 19 incorrect referrals or a number-needed-to-refer of 20). Ang-2 = angiopoietin-2; IL-8 = interleukin-8; LqSOFA = Liverpool quick Sequential Organ Failure Assessment.

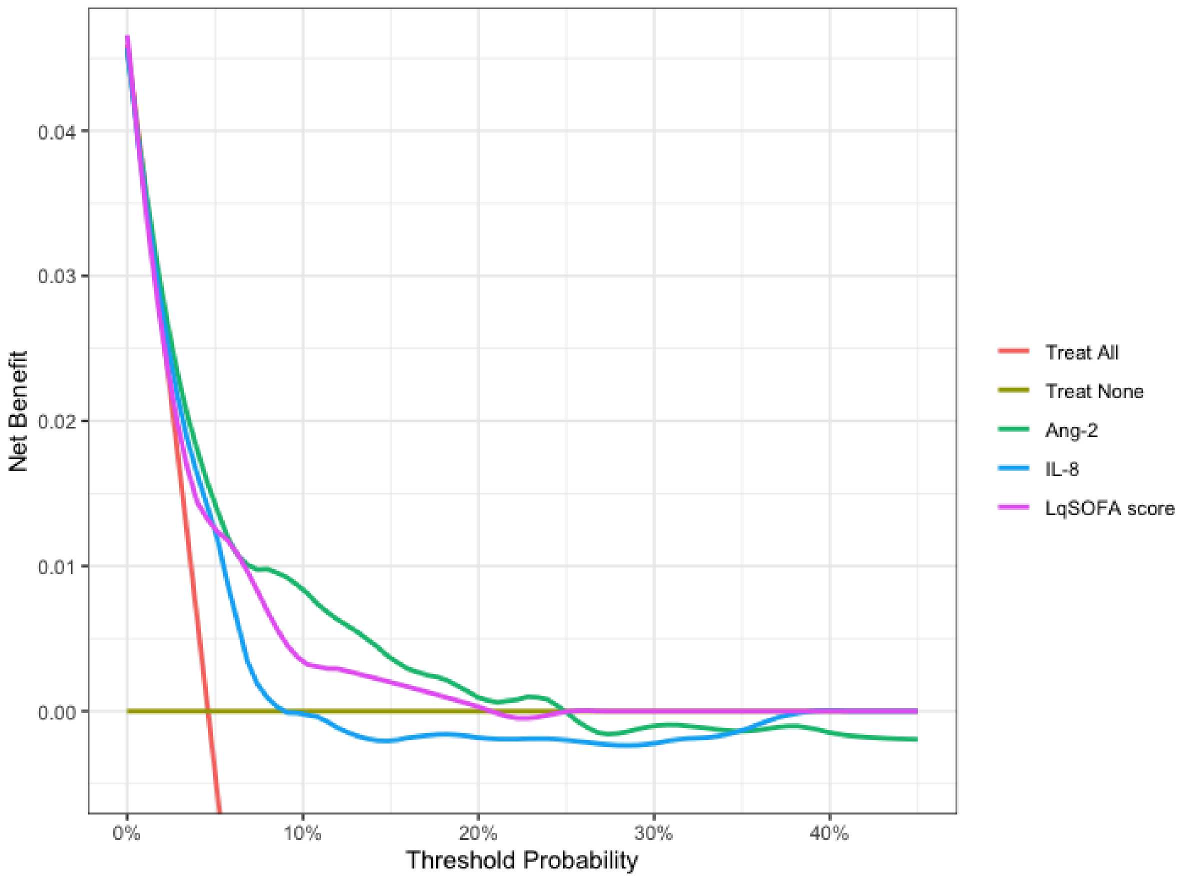

**Supplementary Table 8. Predicted classifications at different referral thresholds using the LqSOFA score and Ang-2.** A referral threshold of 5% reflects a management strategy whereby any child with a predicted probability of requiring oxygen ≥ 5% is referred. *LqSOFA scores converted to predicted probabilities to facilitate comparison with Ang-2; referral thresholds for the LqSOFA score approximate to the following predicted probabilities: 1% ≈ ≥ 0; 5% ≈ ≥ 1; 20% ≈ ≥ 2; 40% ≈ ≥ 3.

| **Referral threshold** | **Sensitivity** (95% CI) | **Specificity** (95% CI) | **Negative Predictive Value** (95% CI) | **Positive Predictive Value** (95% CI) | **Negative Likelihood Ratio** (95% CI) | **Positive Likelihood Ratio** (95% CI) | **Cases referred** (%) | **Cases managed in community** (%) | **Ratio of Incorrect to Correct referrals** | **Ratio of Correct to Incorrect cases managed in community** |
|---|---|---|---|---|---|---|---|---|---|---|
| **LqSOFA score*** | | | | | | | | | | |
| **1%** | 1·00 (NA) | 0·00 (NA) | 1·00 (NA) | 0·05 (0·04-0·07) | 0·00 (NA) | 1·00 (NA) | 900 (100%) | 0 (0%) | 17 to 1 | NA |
| **5%** | 0·79 (0·67-0·89) | 0·83 (0·80-0·85) | 0·99 (0·98-0·99) | 0·21 (0·15-0·26) | 0·25 (0·13-0·40) | 4·56 (3·63-5·54) | 184 (20·4%) | 716 (79·6%) | 4 to 1 | 71 to 1 |
| **20%** | 0·23 (0·13-0·35) | 0·98 (0·97-0·99) | 0·96 (0·94-0·97) | 0·35 (0·18-0·52) | 0·79 (0·67-0·90) | 9·95 (4·62-16·91) | 28 (3·1%) | 872 (96·9%) | 2 to 1 | 22 to 1 |
| **40%** | 0·20 (0·00-0·33) | 0·98 (0·97-1·00) | 0·96 (0·94-0·97) | 0·43 (0·31-1·00) | 0·82 (0·68-1·00) | Inf (NA) | 1 (0·1%) | 899 (99·9%) | 0 to 1 | 18 to 1 |
| **Ang-2** | | | | | | | | | | |
| **1%** | 1·00 (NA) | 0·00 (0·00-0·31) | 1·00 (NA) | 0·05 (0·04-0·07) | 0·00 (NA) | 1·00 (1·00-1·45) | 899 (99·9%) | 1 (0·1%) | 17 to 1 | Inf to 1 |
| **5%** | 0·67 (0·45-0·82) | 0·77 (0·59-0·87) | 0·98 (0·97-0·98) | 0·15 (0·11-0·20) | 0·43 (0·26-0·66) | 3·07 (2·04-4·92) | 225 (25·0%) | 675 (75·0%) | 6 to 1 | 44 to 1 |
| **20%** | 0·24 (0·10-0·42) | 0·97 (0·96-0·99) | 0·96 (0·94-0·97) | 0·35 (0·21-0·49) | 0·78 (0·60-0·92) | 9·65 (5·18-18·48) | 32 (3·6%) | 868 (96·4%) | 2 to 1 | 22 to 1 |
| **40%** | 0·13 (0·03-0·29) | 0·99 (0·99-1·00) | 0·95 (0·94-0·96) | 0·47 (0·15-0·70) | 0·88 (0·72-0·98) | 18·34 (5·72-56·08) | 12 (1·3%) | 888 (98·7%) | 1 to 1 | 20 to 1 |

**Supplementary Figure 5. Net-benefit of Ang-2, the LqSOFA score, and the combination of LqSOFA and Ang-2 to identify children with pneumonia who required supplemental oxygen.** The net-benefit of Ang-2 (pink line) is compared to the LqSOFA (blue line), the combination of Ang-2 and LqSOFA (green line) IL-8 (blue line), and a "refer-all" (red line) and "refer-none" (brown line) approach. A threshold probability of 5% is equivalent to a management strategy in which any child with a predicted risk of supplemental oxygen requirement ≥ 5% is referred (i.e., a scenario where the value of one correct referral is equivalent to 19 incorrect referrals or a number-needed-to-refer of 20). Ang-2 = angiopoietin-2; LqSOFA = Liverpool quick Sequential Organ Failure Assessment.

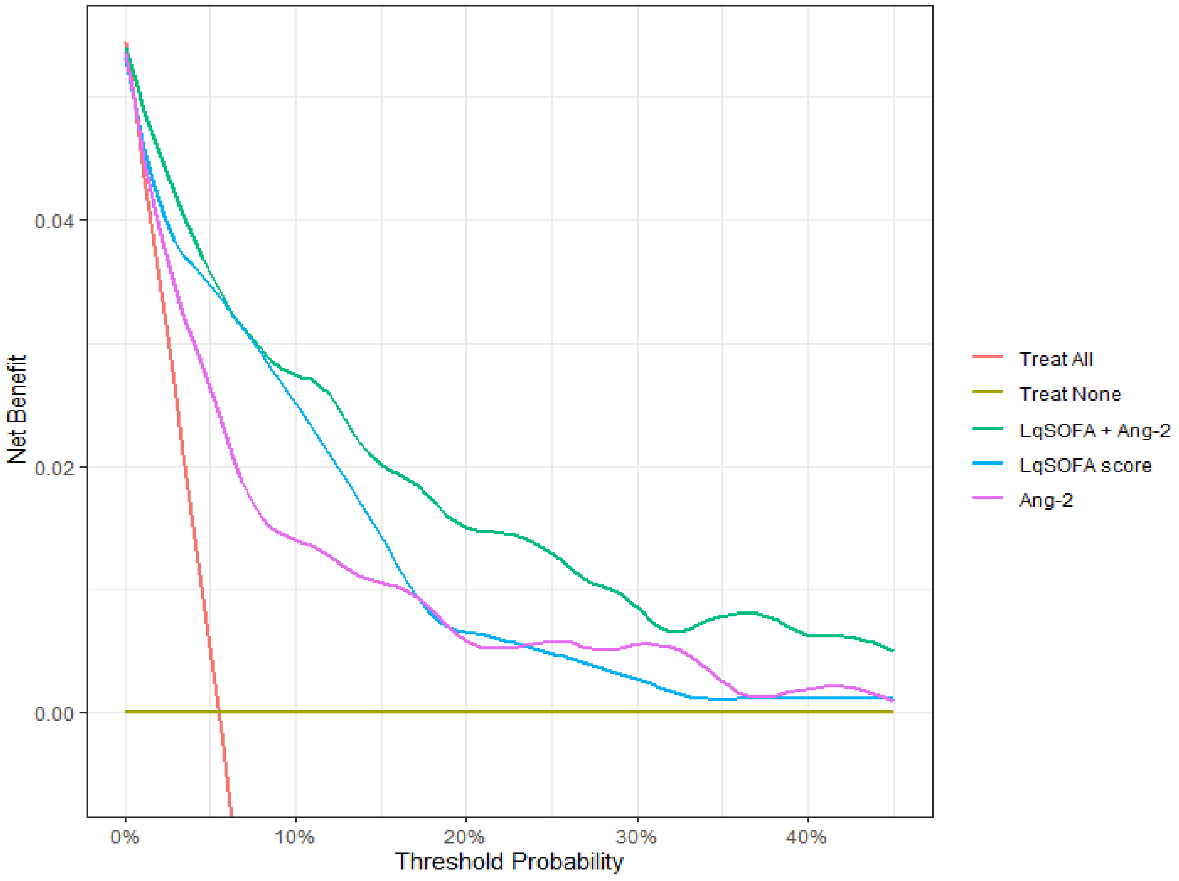

**Supplementary Table 9**. **Comparison of presentations with and without a serum sample available for biomarker analysis.** Presentations without missingness for primary outcome analysed. [a]Respiratory distress defined as head bobbing, tracheal tug, grunting and/or chest indrawing; [b]abnormal chest auscultation defined as crepitations and/or wheeze; [c]rectal temperature converted to axillary temperature for neonates and infants. [†]Age-adjusted z-scores were calculated using the R package *z scorer*. For children admitted to the clinic, weight was measured at the time of presentation (seca scale; precision ± 5g for neonates or ± 50g after birth). All other anthropometric data were captured during routine visits and were eligible for inclusion in these analyses according to the following window periods: height ≤ 28 days; MUAC ≤ 28 days without intercurrent admission; weight ≤ 14 days without intercurrent admission. [*]Missing data: gestation = 3; birthweight = 7; comorbidity = 5; symptom duration = 8; fever = 1; abnormal lung auscultation = 23; lung crepitations = 27; wheeze = 36; heart rate = 5; respiratory rate = 2; temperature = 1; oxygen saturation = 187; capillary refill time = 100; mental status = 18; WLZ = 59 WAZ = 55; MAZ = 135; LAZ = 5. IQR = interquartile range; MUAC = mid-upper arm circumference.

| **Characteristic** | **Overall** N = 900[1] | **Serum sample available** | | **p-value[2]** |
|---|---|---|---|---|
| | | **No** N = 256[1] | **Yes** N = 900[1] | |
| **Demographics** | | | | |
| Age (months) | 10·3 (5·9, 15·6) | 9·4 (5·7, 13·9) | 10·7 (6·0, 16·3) | 0·022 |
| Male sex | 605 / 1,156 (52%) | 149 / 256 (58%) | 456 / 900 (51%) | 0·033 |
| **Birth history** | | | | |
| Gestation (weeks)[*] | 39·1 (38·1, 40·0) | 39·1 (37·6, 40·0) | 39·1 (38·1, 40·0) | 0·077 |
| Birthweight (kg)[*] | 2·9 (2·6, 3·2) | 2·8 (2·5, 3·2) | 2·9 (2·6, 3·2) | 0·40 |
| **Previous medical history** | | | | |
| Number of previous illness visits | 4·0 (2·0, 7·0) | 5·0 (3·0, 7·0) | 4·0 (2·0, 7·0) | 0·059 |
| Time since last illness visit (days) | 35·5 (7·0, 96·2) | 12·5 (3·0, 47·2) | 45·0 (15·0, 106·2) | <0·001 |
| Known comorbidity[*] | 17 / 1,151 (1·5%) | 6 / 255 (2·4%) | 11 / 896 (1·2%) | 0·20 |
| **History of current illness** | | | | |
| Duration of symptoms (days)[*] | 3·0 (2·0, 5·0) | 3·0 (2·0, 6·0) | 3·0 (2·0, 5·0) | 0·70 |

| Characteristic | Overall<br>N = 900[1] | Serum sample available | | p-value[2] |
|---|---|---|---|---|
| | | No<br>N = 256[1] | Yes<br>N = 900[1] | |
| Antibiotics prior to presentation | 79 / 1,156 (6·8%) | 54 / 256 (21%) | 25 / 900 (2·8%) | <0·001 |
| **Presenting symptoms and signs** | | | | |
| Fever[*] | 983 / 1,155 (85%) | 203 / 255 (80%) | 780 / 900 (87%) | 0·005 |
| Cough | 1,148 / 1,156 (99%) | 253 / 256 (99%) | 895 / 900 (99%) | 0·40 |
| Respiratory distress[a] | 403 / 1,156 (35%) | 125 / 256 (49%) | 278 / 900 (31%) | <0·001 |
| Head bobbing | 46 / 1,156 (4·0%) | 13 / 256 (5·1%) | 33 / 900 (3·7%) | 0·30 |
| Tracheal tug | 113 / 1,156 (9·8%) | 37 / 256 (14%) | 76 / 900 (8·4%) | 0·004 |
| Grunting | 21 / 1,156 (1·8%) | 7 / 256 (2·7%) | 14 / 900 (1·6%) | 0·30 |
| Chest indrawing | 395 / 1,156 (34%) | 122 / 256 (48%) | 273 / 900 (30%) | <0·001 |
| Abnormal lung auscultation[b*] | 916 / 1,133 (81%) | 197 / 246 (80%) | 719 / 887 (81%) | 0·70 |
| Crepitations[*] | 833 / 1,129 (74%) | 174 / 244 (71%) | 659 / 885 (74%) | 0·30 |
| Wheeze[*] | 432 / 1,120 (39%) | 94 / 242 (39%) | 338 / 878 (38%) | >0·9 |
| **Vital signs** | | | | |
| Heart rate (bpm) [*] | | | | |
| Neonate | 150·0 (143·8, 161·0) | 159·0 (147·5, 168·5) | 149·0 (143·8, 160·0) | 0·40 |
| Infant | 140·0 (130·0, 148·0) | 140·0 (130·0, 150·0) | 140·0 (130·0, 148·0) | 0·30 |
| Child | 132·0 (124·0, 140·0) | 136·0 (126·0, 148·0) | 132·0 (124·0, 140·0) | 0·024 |
| Respiratory rate (bpm) [*] | | | | |
| Neonate | 64·0 (57·5, 71·5) | 52·0 (42·5, 58·2) | 67·0 (63·5, 77·0) | 0·050 |

| Characteristic | Overall N = 900[1] | Serum sample available | | p-value[2] |
|---|---|---|---|---|
| | | No N = 256[1] | Yes N = 900[1] | |
| Infant | 56·0 (54·0, 60·0) | 56·0 (52·0, 60·0) | 58·0 (54·0, 60·8) | <0·001 |
| Child | 50·0 (46·0, 56·0) | 52·0 (47·0, 58·0) | 50·0 (46·0, 56·0) | 0·054 |
| Axillary temperature (°C)[c*] | 37·1 (36·4, 37·8) | 37·3 (36·4, 37·8) | 37·1 (36·4, 37·7) | 0·064 |
| Oxygen saturation (%)[*] | 94·0 (92·0, 96·0) | 94·0 (92·0, 95·5) | 95·0 (93·0, 96·0) | <0·001 |
| Capillary refill time > 2 secs[*] | 18 / 1,056 (1·7%) | 6 / 219 (2·7%) | 12 / 837 (1·4%) | 0·20 |
| Not alert[*] | 183 / 1,138 (16%) | 67 / 250 (27%) | 116 / 888 (13%) | <0·001 |
| **Anthropometrics** | | | | |
| Weight-for-length z-score (WLZ)[*†] | -0·2 (-0·9, 0·6) | -0·2 (-0·8, 0·8) | -0·2 (-0·9, 0·6) | 0·30 |
| Weight-for-age z-score (WAZ)[*†] | -1·0 (-1·9, -0·4) | -1·0 (-1·9, -0·3) | -1·0 (-1·9, -0·4) | 0·80 |
| MUAC-for-age z-score (MAZ)[*†] | 0·1 (-0·6, 0·7) | 0·3 (-0·5, 0·9) | 0·1 (-0·6, 0·7) | 0·025 |
| Length-for-age z-score (LAZ)[*†] | -1·6 (-2·5, -0·9) | -1·6 (-2·4, -0·9) | -1·6 (-2·5, -0·9) | 0·90 |
| **Primary outcome** | | | | |
| Received supplemental oxygen | 80 / 1,156 (6·9%) | 31 / 256 (12%) | 49 / 900 (5·4%) | <0·001 |

[1]Median (IQR); n / N (%)

[2]Wilcoxon rank sum test; Pearson's Chi-squared test; Fisher's exact test

**Supplementary Table 10. Discrimination of the LqSOFA score and host biomarkers for diagnosis of severe pneumonia, assuming all presentations with missing baseline $SpO_2$ were not hypoxic.** Analysis of 905 presentations, 32 of which met the primary outcome. Only the five top-performing biomarkers from the primary analysis were evaluated for the secondary diagnostic and prognostic outcomes. Ang-2 = angiopoietin-2; AUC = area under the receiver operating characteristic curve; CI = confidence interval; IL-1ra = interleukin-1 receptor antagonist; IL-8 = interleukin-8; LqSOFA = Liverpool quick Sequential Organ Failure Assessment; PCT = procalcitonin; sFlt-1 = soluble fms-like tyrosine kinase-1.

| **Predictor** | **AUC** (95% CI) |
|---|---|
| **LqSOFA** | 0·84 (0·77-0·91) |
| **Ang-2** | 0·75 (0·65-0·85) |
| **sFlt-1** | 0·72 (0·63-0·81) |
| **IL-1ra** | 0·72 (0·61-0·82) |
| **IL-8** | 0·70 (0·62-0·77) |
| **PCT** | 0·69 (0·60-0·78) |

**Supplementary Table 11. Ability of the LqSOFA score and host biomarkers to discriminate children who required supplemental oxygen, excluding presentations on a Saturday or Sunday**. Analysis of 696 presentations, 34 of which met the primary outcome. Ang-1 = angiopoietin-1; Ang-2 = angiopoietin-2; AUC = area under the receiver operating characteristic curve; CHI3L1 = chitinase-3-like protein-1; CI = confidence interval; CRP = C-reactive protein; IL-1ra = interleukin-1 receptor antagonist; IL-6 = interleukin-6; IL-8 = interleukin-8; IL-10 = interleukin-10; IP-10 = interferon-gamma-inducible protein-10; LqSOFA = Liverpool quick Sequential Organ Failure Assessment; PCT = procalcitonin; sFlt-1 = soluble fms-like tyrosine kinase-1; sTNFR-1 = soluble tumour necrosis factor receptor-1; sTREM-1 = soluble triggering receptor expressed on myeloid cells-1.

| **Predictor** | **AUC** (95% CI) |
|---|---|
| **LqSOFA** | 0·82 (0·75-0·90) |
| **Ang-2** | 0·81 (0·75-0·88) |
| **IL-8** | 0·70 (0·62-0·79) |
| **PCT** | 0·67 (0·57-0·76) |
| **sFlt-1** | 0·66 (0·56-0·76) |
| **IL-6** | 0·64 (0·53-0·74) |
| **IL-1ra** | 0·63 (0·52-0·74) |
| **sTNFR-1** | 0·59 (0·49-0·69) |
| **IL-10** | 0·56 (0·46-0·67) |
| **Ang-1** | 0·56 (0·45-0·66) |
| **sTREM-1** | 0·55 (0·47-0·63) |
| **CRP** | 0·55 (0·44-0·66) |
| **IP-10** | 0·54 (0·44-0·64) |
| **CHI3L1** | 0·53 (0·42-0·64) |
